# Oral rabies vaccine baiting and targeted removal strategies for dog-mediated rabies outbreak control: a modelling framework for outbreak management

**DOI:** 10.1101/2025.10.26.25338851

**Authors:** Guan Tong, Rayson Bock Hing Lim, Shi Hui Jin, Kelvin Ho, Alwyn Tan, Malcolm Soh, Wendy Sng, A Janhavi, Gregory Gan, Ma Pei, Nigel Lim, Jue Tao Lim, Tze-Hoong Chua, Borame L Dickens

**Affiliations:** Saw Swee Hock School of Public Health, National University of Singapore and National University Health System; Animal and Veterinary Service, National Parks Board, Singapore; Lee Kong Chian School of Medicine, Nanyang Technological University

**Keywords:** Agent-based model, Importation, Rabies, Vaccination, Zoonosis

## Abstract

**Background:** Support for ongoing regional rabies elimination efforts in dogs and reducing cross border transmission is vital to prevent outbreaks in areas that have previously achieved elimination. However, there are limited studies to estimate their potential use as preventive measures. We explore the effectiveness of two control interventions, oral rabies vaccination (ORV) and targeted ring removal (TRR).

**Methods:** A spatio-temporal compartmental microsimulation model of 2200 free-roaming dogs (FRDs) was fitted to surveillance data with one-time, once-per-year and once-per-month rabies importation events modelled. Based on estimated dog biting rates, seven intervention strategies of TRR and ORV were explored to assess transmission risk reduction in 10,000 model runs.

**Results:** In the absence of intervention, median human infections over 50 months were 0 (IQR: 0–8), 1 (0–11) and 16 (5–29) for one-time, once-per-year, and once-per-month importation events, with 23.5%, 63.1%, and 99.97% of simulations estimating at least one human case. For one-time and annual importation, 50% dog population TRR reduced this to 6.9% and 29.6%, and 90% ORV coverage down further to 5.0% and 23.5%. Under monthly importation, both TRR and ORV did not substantially reduce the number of simulations without infection but did reduce the number of infections down to 4 (0–12) and 3 (0–8).

**Conclusions:** ORV can be successfully utilised for dog-mediated rabies outbreak control in elimination settings but at higher importation rates, may only reduce outbreak size. High TRR rates are required for an equivalent impact which may not be feasible. Additional strong border surveillance and vaccine stockpiling is recommended for outbreak risk mitigation.

## 1. Introduction

Rabies, a viral zoonotic disease caused by the *Lyssavirus*, is transmitted through the saliva of infected animals via mucosal contact or through bites and scratches, which consists of 99% of human rabies cases^1^. This disease results in acute encephalitis, which is invariably fatal once the clinical symptoms appear. According to the World Health Organisation (WHO), rabies causes tens of thousands of human deaths annually, predominantly from dogs, with over 35,000 (58%) of the nearly 60,000 yearly deaths occurring in Asia^2^. Among Southeast Asian countries, nine out of eleven member states in the Southeast Asia region remain endemic for rabies, including Thailand, Philippines and Indonesia^3,4^. In Malaysia, rabies was eradicated in 1999 but has recurred since 2015. These countries report varying incidence^5^ (0.1 to 117.2 per 100,000 people). Singapore has maintained a rabies-free status since 1953^6,7^ yet is at risk of rabies introduction, along with Brunei, due to their geographical adjacency to rabies endemic countries and pet smuggling activities^8^. Overall, concerns of rabies spread across the region have been raised^9^.

Mass vaccination of pet or owned dogs may be done parenterally but when dogs are free-roaming and challenging to capture, alternative control methods need to be considered. Oral rabies vaccination (ORV), recommended by the WHO^10^, has been utilised widely in Europe^11^ and America^12^ to target wild or free-roaming populations of mammalian species susceptible to rabies. Using a genetically stable and apathogenic live-modified rabies vaccine strains such as SAG2 or SPBN GASGAS at high concentrations^13^, ORV had been proven to be highly effective^11,14,15^. For example, SAG2 induced a 79% seroconversion rate in jackals^11^, while SPBN GASGAS induced ORV immune responses in dogs that were not significantly different from those achieved with parenteral vaccination^15^. These are deployed using coating formulas such as palm, coconut or paraffin mixed with fishmeal or other attractants^16^. ORV pilots and trials are ongoing within the region in Indonesia^17^, Thailand^18^, Philippines^19^ and Cambodia^20^. Other rabies control methods include targeted removal, which was utilised in 2009 and 2015 in Bali in response to rabies outbreaks^21^. On the other hand, mass random removal can be counterproductive to disease control as it may lead to unintended consequences such as changes in behaviour (movement, dispersal, and breeding), which may exacerbate disease transmission or exposure. Moreover, the efficacies of depopulation depend on various factors, such as the removal rate and homogeneity in removal implementation^22,23^. Instead, a more effective strategy could be targeted removal within pre-specified radius (i.e. 1km) of dog sightings where dog sterilisation, rehabilitation and rehoming or placement into safe isolated settings could be conducted, although evidence of potential rabies exposure may result in a mixed removal approach being adopted as the disease is incurable and fatal.

As part of a global strategic plan named Zero by 30 by the World Health Organisation, which aims to eliminate dog-mediated human rabies deaths by 2030, control efforts are ramping up^24^. Our study aims to explore the use of preventative control measures by modelling rabies transmission in an urban setting, focusing on the impact of two key strategies: Targeted Ring Removal (TRR) of free-roaming dogs (FRDs) upon identification of a case and ORV. To assist in evidence-based policymaking and scale up trials to mass deployment of ORV or TRR, we ran simulations and explored their estimated effectiveness across a range of reported parameters.

## 2. Methods

### 2.1 Geospatial location and contact matrix of free-roaming dogs

Our study simulated scenarios within a Singaporean context, utilizing multiple datasets on the country’s dog population dynamics sourced from the Animal & Veterinary Service (AVS), a cluster of the National Parks Board of Singapore (NParks) through its Trap-Neuter-Rehome/Release-Manage programme and Community Animal Management teams^7,25^. These datasets reflect the national approach to managing free-roaming dogs (FRDs), which is grounded in the TNRM framework. Singapore has implemented the Trap-Neuter-Release/Rehome-Manage (TNRM) Programme as part of its long-term, sustainable strategy for controlling the free-roaming dog population. Under this programme, free-roaming dogs are humanely trapped, sterilised, microchipped for traceability and rehomed where possible, or rabies-vaccinated for herd immunity and released to live out their lives naturally. The TNRM programme involves close collaboration between the Government, animal welfare groups, veterinarians and the community. The geospatial dataset comprised of 651 unique home territory locations of FRDs observed in the country. In our simulation, we assumed that the total FRD population size is 2200, consisting of (i) unowned, free-roaming individuals characterised by low human interaction (i.e. feeding), and (ii) semi-owned, free-roaming individuals (community dogs) characterised by higher human interaction (i.e. feeding, guard dogs for semi-urban/rural premises)^26,27^.The FRD geospatial distribution were estimated by extrapolating the location information from the observed home territories for the additional FRDs, assuming they stochastically resided within 300 meters of empirical FRD locations^27^.

An *N* · *N* distance matrix, *M* = (*m_ij_*), was generated using the geospatial locations of the dogs, where *m_ij_*represents the distance in metres between dog *i* and *j*. Dogs within 100 meters of each other (i.e., *m_ij_*≤100) were assumed to belong to the same pack, which usually consisted of 3–8 individual dogs^25,28^.

The daily contact matrix, ***P*** = (*p_ij_*), was further derived from the distance matrix *M*, with *p_ij_*representing the probability of contact on a given day between dog *i* and □29. Specifically, we assumed *p_ij_*∼U(0.94,1) for dogs within the same dog pack, while for others we set

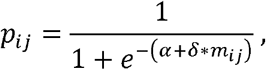

where *α* and *δ* are kernel parameters determing daily contact probability^29^ (Table 1).

**Table 1.** Parameters used within the SEIR model.

| Variable | Value, range or distribution | Definition |
| --- | --- | --- |
| $\alpha$ | 1.6567 | Distance kernel parameter (intercept) describing contact<sup>29</sup> |
| $\delta$ | -0.0159 | Shape parameter (slope) describing contact<sup>29</sup> |
| **M** | $(m_{ij})$ | Distance matrix, with entry $m_{ij}$ representing distance between dog $i$ and $j$ |
| **P** | $(p_{ij})$ | Daily contact matrix, with $p_{ij}$ representing the probability of contact between dog $i$ and $j$ |
| **Q** | $(q_{ij})$ | Daily biting matrix, with $q_{ij}$ representing the probability of dog $i$ being bitten by dog $j$ |
| $b_{ij}$ | U(0.8,0.95) within dog packs,<br>U(0.6,0.8) between dog packs | Probability of dog $j$ biting dog $i$ by during a contact event<sup>29</sup> |
| $\beta_{ij}$ | $q_{ij}v$ | Rate of exposure to rabies for susceptible dog $i$ from rabid dog $j$ |
| $v$ | PERT distribution with support [0.45, 0.52] and mode 0.49 | Probability of contracting rabies after being bitten by a rabid dog<sup>29</sup> |
| $\lambda$ | $\frac{1}{3.1}$ | Rate of removal of dogs due to rabies<sup>30</sup> |
| $\gamma$ | $\frac{1}{22.3}$ | Daily rate of transition from incubation to infectious state for dogs<sup>30</sup> |
| $r$ | 0.001 | Replacement rate of dogs based on available living area (assumed) |

### 2.2 Probability of biting and presence of super-spreaders

The probability of dog *i* being bitten by dog *j, b_ij_*during contact events was modelled as

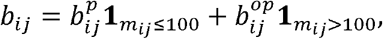

where *m_ij_*is the distance between the two dogs, 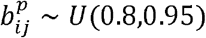 and 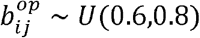 are random variables indicating differentiated chances of biting events for dogs within the same pack and from different packs, respectively.

To account for heterogeneity in FRD behaviour, we designated approximately 4% of the population as potential superspreaders (𝒮), assigning them a tenfold higher contact rate compared to the general population, reflecting their increased likelihood of contacting other dogs^31^. This amplification resulted in approximately 3% of the population interacting with at least 10 other FRDs and up to 30 per day.

In addition, the probability of dog *i* being bitten by dog *j, q_ij_*, was estimated based on a previous methodology^29^ as

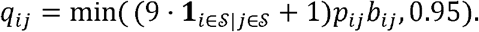

This formed the biting matrix, *Q* = (*q_ij_*), enabling us to construct a network of the 2200 dogs, with nodes representing individual dogs and edges weighted by their daily biting probabilities, *q_ij_*.

### 2.3 SEIR transmission model

An agent-based, spatio-temporal Susceptible-Exposed-Infectious-Removed (SEIR) compartmental model was employed to model rabies transmission among the 2200 dogs. The infection status of each FRD at time *t* was stored and transition probabilities between compartments calculated as

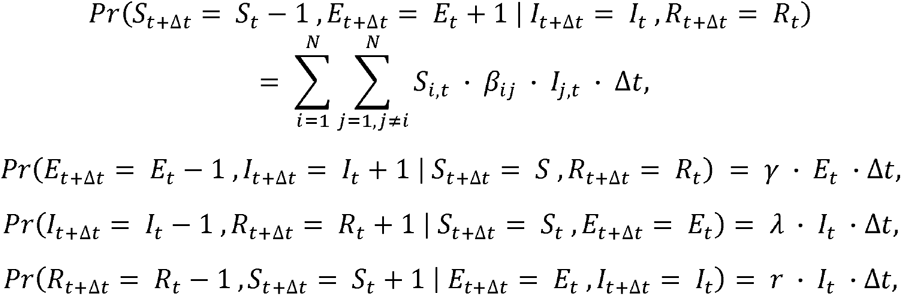

where *β_ij_*-*q_ij_v* is the rate of exposure for a susceptible dog *i* from rabid dog *j*, and *v* the probability of contracting rabies after being bitten by a rabid dog. Dog replacement was assumed upon the death of a dog, in which a new-born, abandoned, or provisioned dog with the same geospatial location and contact/biting probabilities could be added at a replacement rate of. Please refer to Table 1 for details of all the model parameters.

### 2.4 Dog-to-human transmission

The human population was classified into two risk groups based on their contact rates with FRDs. The high-risk group, including FRD feeders (including caregivers from animal welfare groups), children aged 5 to 14, veterinarians, and the elderly, comprised approximately 20% of the Singaporean population^32–34^. The number of humans bitten by rabid dog *i* per day was then simulated from

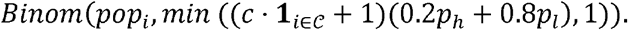

Here, *pop_i_* is the number of humans residing within 520 meters of the dog’s home territory^25^, *c* ∼ *U*(4,9) the increased biting probability for community dogs (𝒞), and *p_h_* and *p_i_* the daily dog-biting probabilities for high- and low-risk human groups, assumed to be 17.60 and 14.99 per 100,000 individuals per day, respectively^35^. Overall rabies exposure in the human population was then quantified by summing the total number of estimated rabid dog bites on humans.

### 2.5 Interventions against rabies

Two intervention methods were considered in this study, including TRR and ORV, to reduce the number of susceptible FRDs and interrupt disease transmission. TRR was conducted through rabid FRD detection and then by randomly removing FRDs whose home territories were located within a one-kilometre radius of detected cases until a pre-specified proportion of the population was removed. Community dogs were assumed to be more likely to be detected than other FRDs, with their detection probability inflated by a factor between five and ten. For the ORV scenario, however, a fixed number of susceptible FRDs were transitioned to a non-infectious group at the end of each month, as these dogs were protected by vaccines and would not become infectious.

To evaluate both measures’ effectiveness in mitigating dog-to-human transmission risk, we proposed the following strategies:

S1: Scenario with no interventions,

S2: TRR of (a) 10%, (b) 20%, or (c) 50% of FRDs, and

S3: ORV campaigns to immunize 50% (a) or 90% (b) of FRDs over six months, or 50% (c) or 90% (d) of FRDs over three months.

The estimated number of rabid dogs and human infections for each strategy was compared to the baseline scenario (S1), in which we did not remove or vaccinate any FRDs. All FRDs were susceptible at the start of the simulation and the outbreak was initiated by an importation event, defined as a secondary local FRD infection resulting from an importation event. This was modelled by changing the status of a randomly-selected susceptible FRD to exposed, implying that it had been exposed to rabies. We considered three levels of importation frequencies, including (i) one-time and (ii) an importation event every 12 months (“Once/Year”), and (iii) an importation event every 6 months (“Once/Month”). We ran 10,000 simulations over 50 months (1500 days) for each combination of intervention strategies and importation frequencies. From these, we derived means, medians, interquartile ranges (IQR), and 95% confidence intervals (CIs) for the number of dog and human infections to estimate centrality and spread.

Transmission risks were quantified as the proportion of simulations where outbreaks of different threshold sizes occurred. All analyses and visualizations were performed using R software^36^.

## 3. Results

### 3.1 Geospatial distribution of FRDs

Substantial variation was observed in the contact frequencies among the 2200 FRDs in Singapore. Approximately 3.0% of the population interacting with at least 10 other FRDs and up to 30 per day. Another 12.3% of the population encountered at least five but fewer than 10 FRDs daily, while 40.1% likely to have no interaction any other FRD in a typical day (Figure 1).

**Figure 1.**
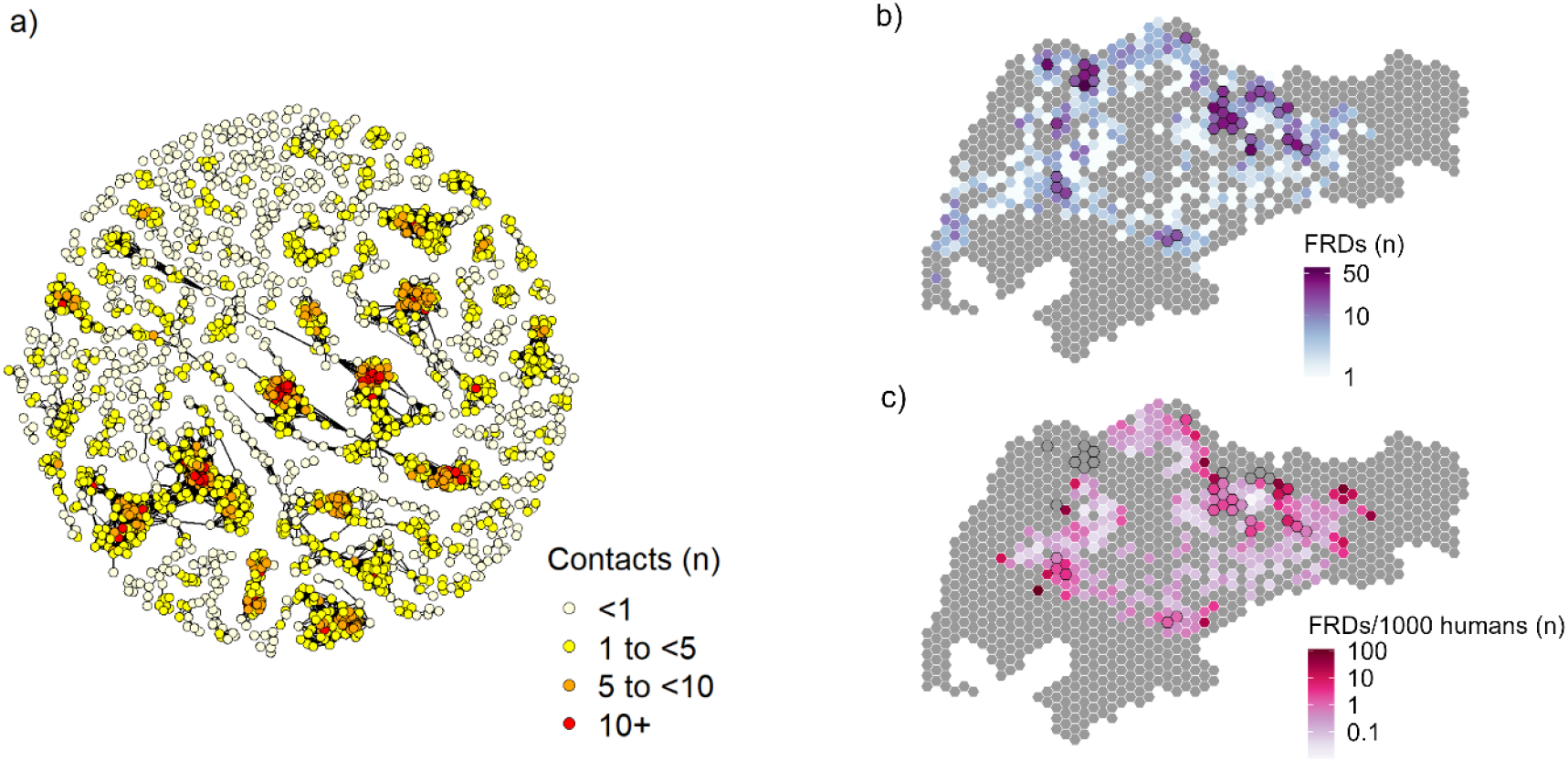
Connectivity and spatial density of the 2200 FDRs: (a) Individual connectivity network of FRDs with clusters of packs, measured by the average number of FRDs contacted per day, (b) Spatial distribution of FRDs in Singapore, (c) Number of FRDs per thousand human residents in each area. Note that ratios in (c) were not calculated for areas without human residents. High-density spatial units, those with over 15 FRDs, are highlighted with a black border in subfigures (b) and (c).

FRDs were also unevenly distributed across the country. Among all spatial units, each covering approximately 0.9km^2^, 9.4% had at most one FRD and 19.5% had no greater than 10. High-density spatial units, defined as those with over 15 FRDs per unit, accounted for 3.2% of the country’s area. These units were mainly concentrated in northern and western Singapore, where clusters of three or more connected high-density units were observed. The highest dog concentration was observed at one spatial unit in the northwest, in which up to 62 FRDs gathered. After adjusting for human population density, central west and central north remained as regions with high FRD populations, with at least one dog per 1000 humans. The northeastern area also emerged as an area with high FRD-to-human ratio, potentially due to low human density. It is also worth noting that some areas with high FRD populations, such as the aforementioned FRD clusters in northwestern Singapore, had no human residents and were therefore excluded from ratio calculations (Figure 1).

### 3.2 Dog-to-dog transmission dynamics

With a one-time importation event, the risk of having at least one active rabid FRD peaked at 26.5% in the eighth month following initial importation but gradually declined to less than 4.9% from month 36 onwards to a minimum of 1.7% by the end of the 50-month period. When the importation frequency increased to once-per-year, intermittent outbreaks were observed, with 83.9% of the 1500 days having over 25% risk of at least one infectious FRDs, and 32.4% of the 1500 days having over 25% risk of at least 2 infectious FRDS, respectively. In the monthly-importation scenario, however, the number of infectious FRDs increased rapidly to 7 (IQR: 1–17) within the first two years and remained constant at approximately 10 (IQR: 3–20) from month 36 onwards (Figure 2).

**Figure 2.**
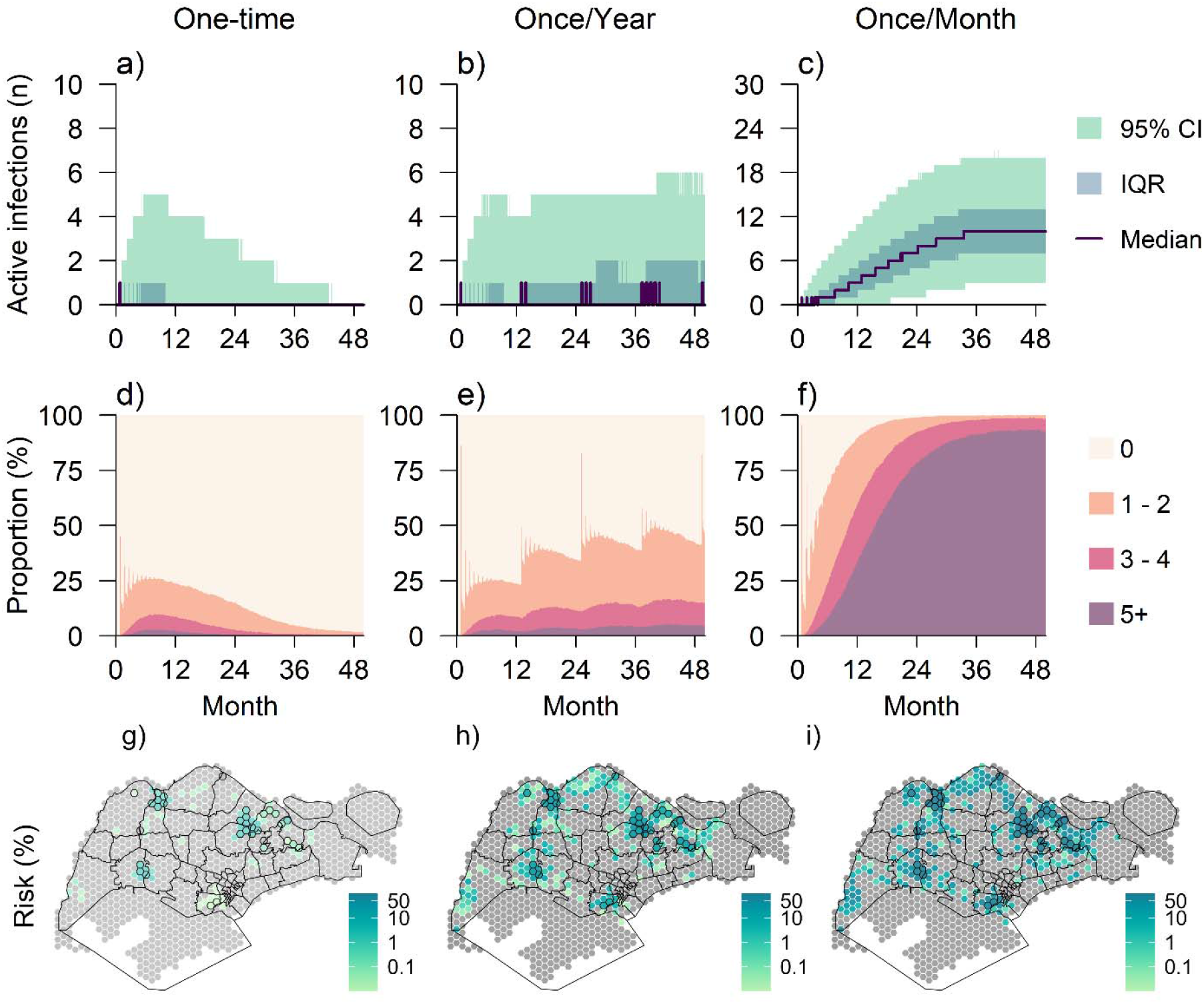
Dog-to-dog transmission dynamics. Row 1 (a–c) shows a time series of infectious FRDs per day, in which the solid lines represent medians, and darker and lighter shades represent the interquartile ranges (IQR) and 95% confidence intervals (CI), respectively. Row 2 (d–f) presents proportions (%) of simulations with zero (‘0’), one to two (‘1–2’), three to nine (‘3–4’), five or more (‘5+’) infectious FRDs in each day over the 50 months. Row 3 (g–i) displays the geospatial distribution of the risk for at least one dog-to-dog transmission event over the 50-month period, in which high-density spatial units (zones with over 15 FRDs) are highlighted with black borders. The three columns correspond to the scenarios of one-time (1), once-per-year (2), and once-per-month (3) importations, respectively.

Under the one-time importation scenario, 45.5% of the high-density spatial units had a 1% or higher of experiencing at least one rabies transmission event between dogs, comprising 66.7% of all such high-risk regions. In the yearly-importation scenario, the average risk in the high-density spatial units rose to 15.8% with 21 units showing a risk of 10% or more. With monthly importation, while 16.8% of the country’s area had over a 10% risk of at least one secondary rabies transmission between FRDs, the proportion rose to 100% among the high-density units, where the risk averaged 90.6% and ranged from 67.5% to 99.9% (Figure 2).

### 3.3 Dog-to-human transmission risk

The risk of a zone having more than one human infection remained below 0.5% over the 50 months with a one-time importation, and increased to a maximum of 1% with yearly importation. When importation occurred monthly, however, only 20 out of the 50 months maintained a risk below 5%, with one month reaching as high as 10% (Figure 3).

**Figure 3.**
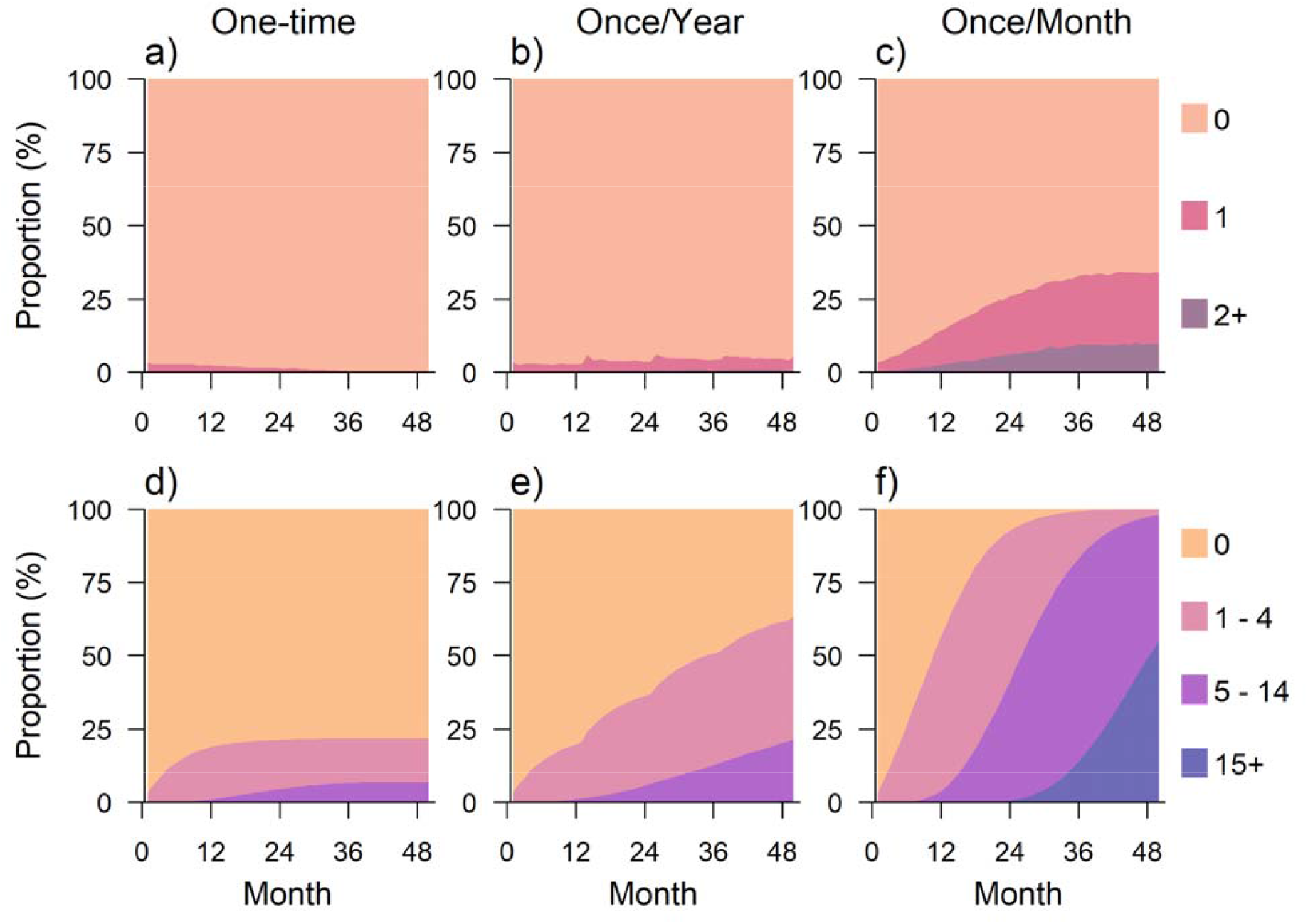
Distributions of simulated human infection counts. Proportions refer to the percentages of the simulated results falling into each category across the 10 000 simulations. The statistics of interest include new infections (Row 1) and cumulative infections (Row 2) across the 50 months, under scenarios of one-time (Column 1), once-per-year (Column 2), and once-per-month (Column 3) importations.

The disparities in the cumulative human cases were also more pronounced. By three years, the probability of having at least one human infection was 21.7%, and that of having five or more cases was 6.6% in the one-time importation scenario. These proportions rose to 60.7% and 11.4% when importation was once-per-year. With once-per-month importation, the chance of having fewer than five infections dropped to 16.9%, while the risk of at least 15 cases occurring was as high as 13.8% (Figure 3).

### 3.4 Effectiveness of interventions

In the no intervention scenario, the median number of human infections over 50 months was 0 (IQR: 0–8), 1 (IQR: 0–11), and 16 (IQR: 5–29) when the importation frequency was one-time, once-per-year, and once per month, respectively, while the risk of at least one human infection occurring over 50 months for the respective three importation frequencies were 23.5%, 63.1%, and 99.97% (Figure 4).

**Figure 4.**
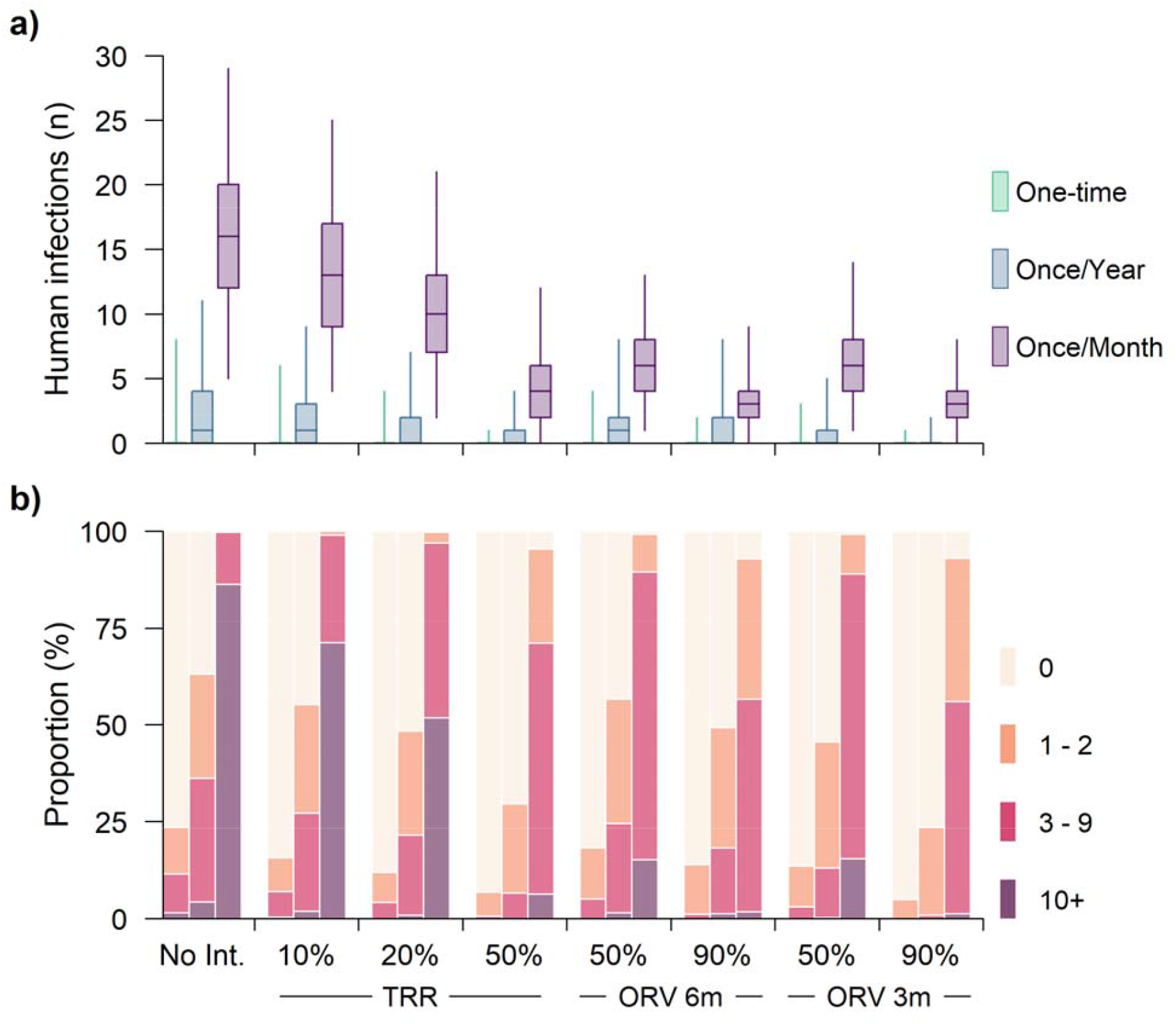
Number of human infections across different intervention strategies: (a) Absolute number (median, IQR, and 95% CI) of human infections which took place in 50 months when different interventions were implemented. Statistics for the one-time, once-per-year, and once-per-month importation scenarios were presented in green, cyan, and purple, respectively. (b) Proportions (%) of simulations with zero (‘0’), one to two (‘1–2’), three to nine (‘3–9’), 10 or more (‘10+’) human infections occurred over the 50-month period. No Int. stands for no interventions, and TRR and ORV stand for targeted ring removal and oral rabies vaccination, respectively. Coverage for the one-time, once-per-year, and once-per-month importation scenarios are displayed from left to right for each strategy.

With a one-time importation event, TRR is estimated to lower the probability of at least one human infection occurring from 23.5% in the no-intervention scenario to 15.8%, 12.0%, and 6.9% with the removal of 10%, 20%, and 50% of FRDs, respectively. In the once-per-year importation scenario, the probability decreased from 63.1% in the no-intervention scenario to 55.3%, 48.4%, and 29.6% with the respective removal rates. Under once-per-month importation, however, this intervention had a limited effect on eliminating dog-to-human transmission, with 99.95%, 99.76%, 95.45% of simulations still showing at least one human infection. Nonetheless, it did decrease the number of human infections to 13 (IQR: 4–25), 10 (IQR: 2–21), and 4 (IQR: 0–12), respectively (Figure 4), compared to 16 (IQR: 5–29) with no intervention.

With a one-time importation event at 50% and 90% ORV coverage, the probability of at least one human infection occurring reduced from 23.5% in the no-intervention scenario to 16.5% and 9.7%, respectively, and from 63.1% with no intervention to 57.9% and 53.6% under once-per-year importation. Similarly, under once-per-month importation, ORV also had limited ability to eliminate human infection, which remained present in 99.16% and 92.86% of the simulations. In scenarios with monthly importation events, human infections dropped from 16 (IQR: 5–29) with no intervention to 6 (IQR: 1–13) when 50% of FRDs were immunized and further to 3 (IQR: 0–9) when 90% of FRDs were immunized within six months. The additional benefits of rapid ORV implementation within 3 months, and not 6, varied across different importation frequencies, which is able to further reduce the probability of at least one human infection occurring down to 13.7% and 5.0% with 50% and 90% coverages of one-time importation, 45.6% and 23.5% of once-per-year importation, while has limited effectiveness under once-per-year importation. (Figure 4).

## 4. Discussion

Our simulation results indicate a high possibility of sustained local rabies transmission following multiple importation events. While a single importation would probably trigger only a small-scale outbreak with no more than five active infections at its peak, more frequent importation events, such as on a monthly basis, could lead to an endemic state with ongoing transmission of up to 10 (IQR: 3–20) active infectious FRDs in the community. The high mobility of FRDs greatly facilitates rabies spread once a dog becomes infectious, especially when a super-spreader is infected which can encounter at least 10 dogs per day. Such outbreaks have been observed in Tunisia^37,38^.

While Singapore remains free from rabies due to strict biosecurity measures, its status is continuously at risk from its proximity to rabies-endemic areas or countries^39^ where cross-border transmission of rabies could occur through dog movement. As a major global hub for trade, transport, and tourism, the country also faces the risk of accidental introduction of rabies through imported animals or travellers^40^. To mitigate such risks, Singapore has implemented a robust and multi-layered regulatory framework for rabies prevention^41^. Pre-import controls require animals to fulfil all conditions of the import licence prior to arrival in Singapore. Depending on the assessed rabies risk of the exporting country, these measures include a minimum residency period of six months, rabies vaccination, and serological testing. At the border, live animals are subject to inspection upon entry, and dogs and cats are quarantined post-arrival by default, with the duration of quarantine based on the exporting country’s rabies status. These stringent controls significantly reduce the likelihood of rabies incursion through imported animals. Given these biosecurity measures, the probability of monthly importation events remains low. However, our simulations highlight the imperative of vigilant border control, particularly in the inspection of incoming dogs that may be incubating rabies— especially from endemic regions—given the disease’s long and often asymptomatic incubation period^42,43^.

Our projection of intervention effectiveness shows that TRR with a moderate removal rate of 10% would only lead to a minor reduction in dog-to-human transmission events, providing limited contributions to curbing rabies transmission. Increasing the removal rate to 20% or 50%, whilst desirable, may be challenging many contexts as FRDs are usually highly mobile, difficult to locate and challenging to capture^28^. By comparison, mass vaccination, achieving 90% immunization of FRDs against rabies, was projected to be at least as effective in averting human infections as removing 50% of FRDs through TRR. With monthly importation, if mass vaccination with 90% coverage is implemented within three months following the initial importation event, only three (IQR: 0–8) individuals are estimated to be infected over 50 months. Similar results were observed with 70% coverage^44^. Additionally, the success of mass vaccination in dog rabies management has been recently demonstrated in Mexico, which eradicated dog mediated rabies in 2019 through vaccinating over 80% of stray dogs in the country^45,46^. All the proposed intervention strategies were estimated to not eliminate dog-mediated rabies with regular importation events of once-per-year or once-per-month. When immunizing 90% of FRDs, it remains likely that at least one human case would occur over the 50-month period following the initial importation event. Vaccine stockpiling is therefore necessary regardless of the effectiveness of ORV.

Our study had several limitations. While the maximum effectiveness of TRR was capped at 50%, this might not reflect real-world performance, particularly when TRR is implemented alongside with sterilisation programmes (i.e. TNRM) which could result in lower FRD replacement rates and reduced disease transmission^47^. Additionally, vaccination coverage may fall below 90%^11^ dependent on bait uptake, and could be challenging to monitor over time. We also assumed a fixed FRD population over the 50-month simulation window with constant replacement from births (i.e. unsterilised populations) whereas dog populations may be more stochastic with diminishing replacement rate, although the degree of annual change is currently not known. Pet abandonment or spikes in birth rates^48,49^ have been documented although the impact of these on FRDs has not been documented. Dog packs may also change in their size and locations from ongoing control efforts and thus contact patterns may also consequently. Heterogeneity in interactions between FRDs, pet dogs, community dog feeders, veterinarians among other groups were not modelled owing to the paucity in data on their contact frequencies. An importation event in our study was defined as a secondary local FRD infection caused by an imported infected dog. While this approach circumvents the complexity of classifying importation sources and determining their connectivity with the local FRD population, it limits our ability to model scenarios involving multiple secondary infections from a single source. Nonetheless, such events are expected to be rare, given the contact patterns among FRDs and other dog populations, and the relatively short infectious period^49^.

Our study examined two intervention strategies, TRR and ORV, for controlling outbreaks in settings currently affected by or at risk of rabies importation. Many countries in Southeast Asia are still endemic for canine rabies though Singapore and Brunei have achieved and maintained freedom from the disease^5,50^. For endemic countries aiming to reduce prevalence or eliminate canine rabies, ORV alongside with robust surveillance and strict enforcement, could be utilised to control and manage the disease. While oral rabies vaccines offer a promising strategy for achieving herd immunity and curbing canine rabies spread in endemic areas, they are generally not suitable for routine use in rabies-free settings. This is because all currently used oral rabies biologics are modified-live, attenuated, or recombinant vaccines, posing a potential risk of residual pathogenicity to non-target species (i.e., wildlife)^51^. Additionally, their use could complicate ongoing surveillance efforts crucial for maintaining rabies-free status. Depending on the accessibility to dog population, mixed-methods vaccination campaigns that involved parenteral vaccination for dogs that are highly accessible (i.e. owned and confined dogs), complemented with oral rabies vaccination for inaccessible dogs (unowned, free-roaming) is recommended to enhance vaccination coverage for canine rabies outbreak control by WOAH and WHO^52^. The implementation of any vaccination or population management programme should specifically target dog populations that are critical for outbreak control or prevention.

This study demonstrates the comparative effectiveness of TRR and ORV strategies for rabies outbreak control and highlights the importance of maintaining strict import controls and robust surveillance systems for preventing rabies incursion. The simulation framework developed here facilitates the identification of at-risk populations for targeted management (i.e. vaccination or removal) and provides evidence-based guidance for effective response to control rabies outbreak.

## Data Availability

All data produced in the present study are available upon reasonable request to the authors

## Supplementary Information

**Figure S1.**
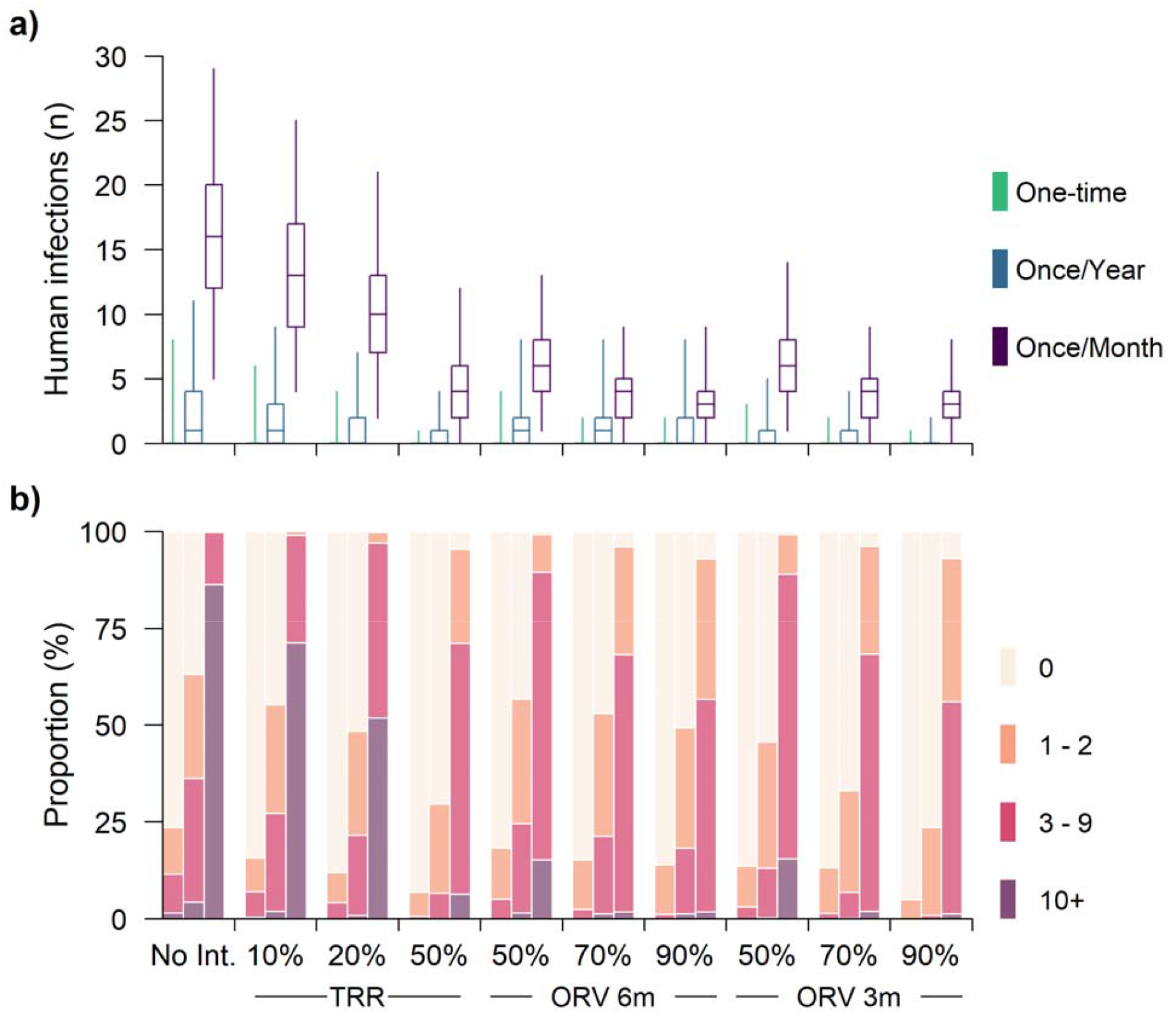
Number of human infections across different intervention strategies with an additional 70% coverage for ORV: (a) Absolute number (median, IQR, and 95% CI) of human infections which took place in 50 months when different interventions were implemented. Statistics for the one-time, once-per-year, and once-per-month importation scenarios were presented in green, cyan, and purple, respectively. (b) Proportions (%) of simulations with zero (‘0’), one to two (‘1–2’), three to nine (‘3–9’), 10 or more (‘10+’) human infections occurred over the 50-month period. No Int. stands for no interventions, and TRR and ORV stand for targeted ring removal and oral rabies vaccination, respectively. Coverage for the one-time, once-per-year, and once-per-month importation scenarios are displayed from left to right for each strategy.

## Notes

### Competing Interest Statement

The authors have declared no competing interest.

### Funding Statement

This work was supported by the National Parks Board Fund [WBS A-8001402-00-00]; PREPARE Ministry of Health [WBS A-8000642-01-00].

